# Underhydration is prevalent across education levels and associated with low intake of water but not sugar-sweetened beverages: A cross-sectional study from the UK

**DOI:** 10.64898/2026.01.28.26344904

**Authors:** Maria Almudena Claassen, Esther Papies

**Affiliations:** Max Planck Institute for Human Development, Center for Adaptive Rationality, Lentzeallee 94, 14195, Berlin, Germany; Behavioural Science Institute, Faculty of Social Sciences, Radboud Universiteit, Nijmegen, Netherlands

**Keywords:** fluid intake, hydration, urine colour, education, water, sugar-sweetened beverages

## Abstract

**Purpose:** Adequate hydration is vital for health, yet many people do not meet fluid recommendations. This study aimed to characterise the role of water and sugar-sweetened beverages in hydration across different levels of socioeconomic status (SES) in the UK.

**Methods:** In a pre-registered cross-sectional study, participants (*N* = 1,112) recalled beverages consumed on the previous day and reported urine colour as an indicator of their hydration status. We analysed water intake (H1), sugar-sweetened beverage (SSB) intake (H2), and SES (education; H3) as predictors of hydration status using stepwise binomial logistic regression adjusted for health, demographic, and lifestyle covariates.

**Results:** Forty percent of participants were classified as underhydrated. Higher water intake was associated with a greater likelihood of adequate hydration: Drinking one extra glass of water per day (250 ml) increased the odds of being adequately hydrated by about 16%. However, SSB intake was not associated with hydration unless intake from other drink sources was held constant. Having a higher versus lower level of education was not significantly associated with hydration status, although finer-grained and income-based analyses suggested modest socioeconomic differences.

**Conclusion:** Water intake—rather than SSB intake—is the primary correlate of adequate hydration in this UK sample. Public health initiatives should emphasise the importance of water for hydration, invest in ways to make water more appealing, and promote the use of urine colour as a marker of hydration status.

---

Adequate hydration is fundamental for human health. A growing body of evidence shows that underhydration is associated with a higher risk of chronic disease, accelerated biological ageing, and declines in cognitive performance [1–3]. Hydration reflects the balance between fluid intake and fluid loss. Public health recommendations therefore emphasise regular intake of sufficient fluids from food and drinks—approximately 2 litres per day for women and 2.5 litres per day for men [4]. Yet population-level data indicate that these recommendations are frequently unmet: A review of fluid intake across 13 countries found that around 50% of women and 60% of men consumed less water from fluids than advised [5].

Biomarker studies further corroborate that inadequate hydration is widespread. For example, Malisova [6] observed that nearly one in five adults they followed over 7 consecutive days was dehydrated. Compared with adequately hydrated participants, dehydrated participants consumed significantly less fluid, had lower urine volume, and had darker urine colour. Urine colour, which correlates with urine osmolality and urine-specific gravity [7], has been proposed as an adequate and accessible proxy for hydration status both in research and for laypeople [8,9].

Although many studies treat fluid intake as a single, uniform category, recent evidence suggests that the hydration potential of different beverage types differs [10]. Prior work in a UK adult sample found that better hydration status—as indicated by lighter urine colour—was associated with higher intake of water and lower intake of sugar-sweetened beverages (SSBs; [11])^1^. Similar patterns between beverage intake and urine concentration (osmolality) have been observed in children and may partly reflect substitution effects: People who drink more SSBs consume less water [12,13]. Differentiating between beverage types has public health significance not only because understanding hydration is essential for human physiological and functioning, but also because SSB intake carries additional health risks, including obesity and metabolic disease [14].

Socioeconomic status (SES) may also shape patterns of beverage consumption and hydration. Brooks et al. [15] found that adults from racially, ethnically, and socioeconomically disadvantaged backgrounds in the US were more likely to be inadequately hydrated, partly due to drinking less tap water and other non-caloric beverages. Education-related differences in SSB intake have likewise been observed in other contexts, such as Norway [16]. However, the association between intake of beverage types and hydration status across has not yet been examined in the UK population. The current study aims to fill this research gap.

## The current study

This cross-sectional study sought to clarify the associations between water intake, SSB intake, and hydration status in a large UK sample with balanced representation across education levels as a proxy for SES. Drawing on the literature reviewed above, we pre-registered the following hypotheses: (H1) Water intake is positively associated with hydration status, (H2) SSB intake is negatively associated with hydration status, and (H3) higher SES is positively associated with hydration status. All hypotheses were preregistered prior to data collection on the Open Science Framework (https://osf.io/jrwyh/overview?view_only=b51a7b252adc46288a27f3ed46c11c18). All materials, data, and analysis scripts are available there and on Github (https://github.com/maclaassen/hydration). In our analyses we clearly separate confirmatory and exploratory analyses, and all measures and exclusions are reported. Additional analyses are reported in the Supplementary Online Material (SOM); those necessary to contextualise the findings of the current paper are reported in the text as well.

## Method

### Participants

A power analysis based on the association between SSB intake and hydration status reported in a previous study [11] indicated that a sample size of *n* = 200 would provide 90% power to replicate this correlation (see SOM for details). Because we planned comparisons across education levels, we preregistered a minimum of 200 participants from each of six education levels.

Participants living in the UK were recruited via Prolific (*Prolific.co* [17]). We recruited equal numbers of male and female participants. Ninety-one percent of participants completed the survey on Wednesday, 7 September 2022, recalling drinks consumed on the previous day. To improve representation in the lowest education category, we continued data collection until 14 September 2022; however, we did not reach the preregistered target for that group. Temperatures varied across regions of the UK and assessment days (maximum temperatures between 17°C and 26°C); we therefore controlled for variation due to location and assessment day in the analyses. We also excluded submissions made on weekend days (*n* = 3).

Following preregistered exclusion criteria, we excluded *n* = 53 participants who reported being ill (i.e., having a flu/cold or fever, vomiting and/or diarrhoea, or a urinary tract infection), and *n* = 32 participants who reported implausible fluid intakes (< 0.4 litres or > 6 litres). The final sample was *N* = 1112 (see Table 1).

**Table 1.** Sample descriptives by education level: n (%) or Mean (SD)

| Characteristic | Overall<br>*N* = 1,112 | No formal<br>qualifi-<br>cations<br>*N* = 37 | Secondary<br>education<br>*N* = 225 | High school<br>diploma/<br>A-levels<br>*N* = 210 | Technical/<br>community<br>college<br>*N* = 216 | Under-<br>graduate<br>degree<br>*N* = 215 | Graduate/<br>doctorate<br>degree<br>*N* = 209 |
| --- | --- | --- | --- | --- | --- | --- | --- |
| Age |  |  |  |  |  |  |  |
| 18–34 | 391 (35%) | 11 (30%) | 43 (19%) | 103 (49%) | 61 (28%) | 89 (41%) | 84 (40%) |
| 35–54 | 470 (42%) | 6 (16%) | 108 (48%) | 74 (35%) | 95 (44%) | 95 (44%) | 92 (44%) |
| 55–85 | 251 (23%) | 20 (54%) | 74 (33%) | 33 (16%) | 60 (28%) | 31 (14%) | 33 (16%) |
| Gender |  |  |  |  |  |  |  |
| Female | 554 (50%) | 21 (57%) | 113 (50%) | 102 (49%) | 110 (51%) | 98 (46%) | 110 (53%) |
| Male | 547 (49%) | 15 (41%) | 110 (49%) | 104 (50%) | 105 (49%) | 116 (54%) | 97 (47%) |
| Non-binary/<br>other | 8 (0.7%) | 1 (2.7%) | 2 (0.9%) | 3 (1.4%) | 1 (0.5%) | 1 (0.5%) | 0 (0%) |
| Unknown | 3 | 0 | 0 | 1 | 0 | 0 | 2 |
| Income |  |  |  |  |  |  |  |
| Low<br>($< \pounds 40,000$) | 603 (55%) | 30 (83%) | 159 (71%) | 106 (51%) | 138 (64%) | 94 (44%) | 76 (37%) |
| High<br>($\geq \pounds 40,000$) | 500 (45%) | 6 (17%) | 66 (29%) | 101 (49%) | 76 (36%) | 119 (56%) | 132 (63%) |
| Unknown | 9 | 1 | 0 | 3 | 2 | 2 | 1 |
| Hydration<br>status (urine<br>colour) |  |  |  |  |  |  |  |
| Inadequate | 450 (40%) | 23 (62%) | 99 (44%) | 83 (40%) | 100 (46%) | 74 (34%) | 71 (34%) |
| Adequate | 662 (60%) | 14 (38%) | 126 (56%) | 127 (60%) | 116 (54%) | 141 (66%) | 138 (66%) |
| Total fluid<br>intake (l) | 2.64 (1.09) | 2.54 (1.16) | 2.49 (1.08) | 2.65 (1.11) | 2.54 (0.97) | 2.76 (1.12) | 2.78 (1.12) |
| Total water<br>intake (l) | 1.06 (0.99) | 0.77 (1.09) | 0.94 (0.93) | 1.12 (1.03) | 0.90 (0.88) | 1.17 (1.04) | 1.22 (1.03) |
| Total SSB<br>intake (l) | 0.26 (0.54) | 0.23 (0.63) | 0.26 (0.50) | 0.28 (0.53) | 0.25 (0.52) | 0.28 (0.59) | 0.23 (0.52) |
| BMI | 27.1 (5.9) | 26.3 (6.6) | 27.7 (5.7) | 27.1 (6.7) | 27.8 (6.0) | 26.6 (5.4) | 26.3 (5.2) |
| Moderate–<br>intense<br>physical<br>activity (min) | 26 (48) | 17 (33) | 21 (40) | 23 (41) | 27 (49) | 33 (65) | 26 (44) |
| Unknown | 5 | 1 | 1 | 0 | 0 | 1 | 2 |

### Measures and procedure

Participants provided informed consent and completed the online survey at a location of their choosing. They were informed that they would have to recall all drinks they had consumed on the previous day and that they would be asked about their lifestyle and health (e.g., physical activity, food consumption practices, and urine colour).

#### Beverage consumption

Participants were asked to report all drinks consumed on the previous day using an adapted 1-day version of the Liq.In7 [18] (see Fig. S1 in SOM). They reported all beverages consumed in the morning, afternoon, and evening, with prompts such as “before breakfast,” “during lunch,” and “at night” aiding retrieval. For each drink, they reported the drink type, the specific drink, the container from which they drank, the quantity consumed, and whether they added sugar. Participants selected from seven drink types: water, hot drinks, milk drinks, soft/flavoured beverages, diet beverages, alcohol, and other. Each drink type had specific drinks as subcategories. Container types included glass/tumbler, plastic/foam/paper cup; participants estimated how much of the container they consumed (e.g., “half of it,” “all of it”). They also indicated whether they added sugar to their drink and, if so, how much (e.g., “no,” “1 spoon”; see Fig. 1 for an overview of the method). Finally, participants were asked whether they had soup or milk with their cereal, and if so, what size bowl they had used.

**Fig. 1.**
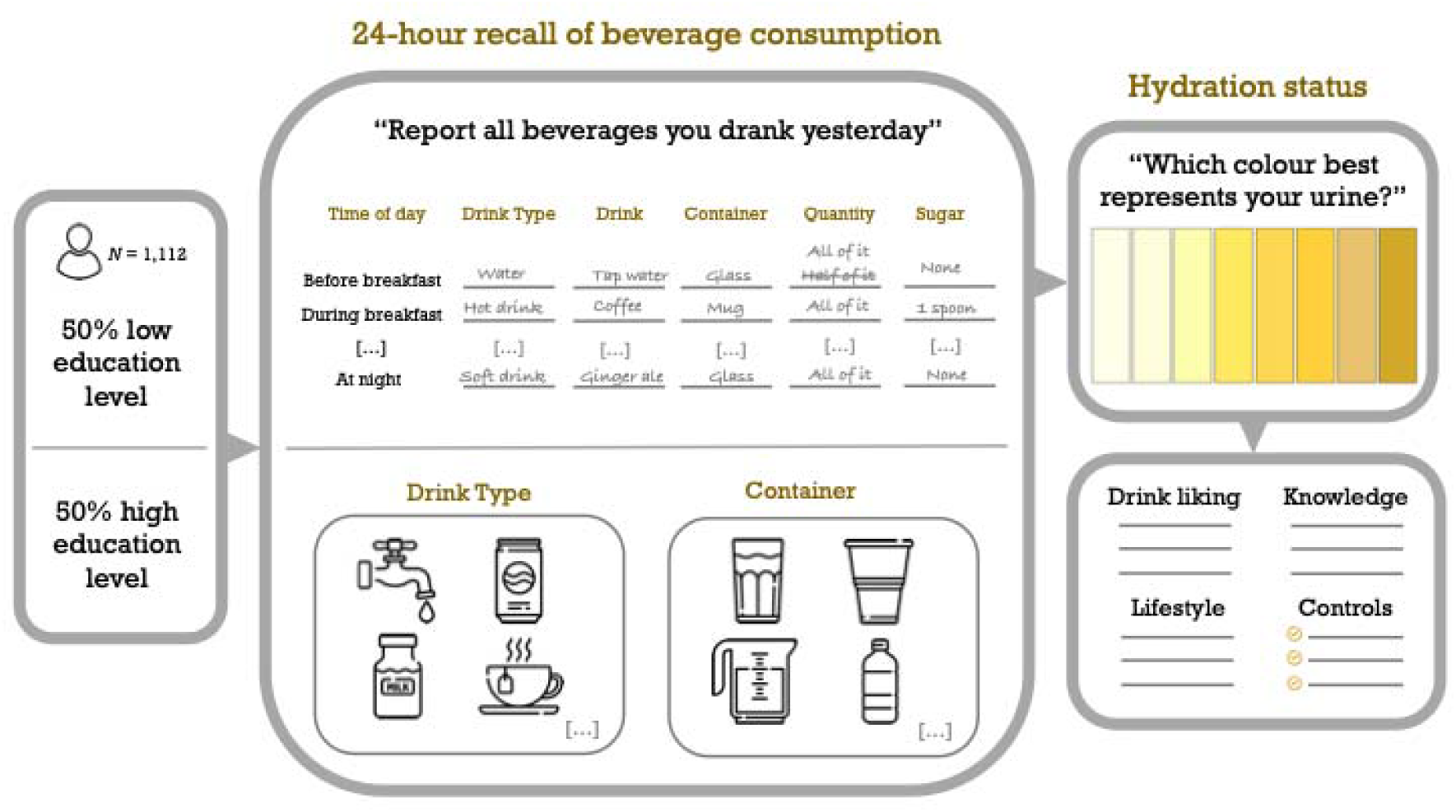
Overview of the methodology.

Total fluid intake was calculated from all reported drinks (including fluid from milk in cereal and from soup). Water intake comprised all drinks in the water category: tap water, water from cooler/fountain, and bottled water (still or carbonated, flavoured or unflavoured). SSB intake comprised all drinks in the soft/flavoured beverages category: carbonated soft drinks, non-carbonated sweet drinks, iced tea, and other ready-to-drink sweetened beverages (excluding energy/sport drinks and packaged fruit and vegetable juices).

#### Hydration status

The primary assessment of hydration status was urine colour on an eight-colour urine chart [19] (see Fig. 2). Participants selected the colour best representing the colour of their urine on the previous day. The three lightest urine colours were defined as indicating adequate hydration (corresponding with a state of euhydration) and the five darkest colours as indicating inadequate hydration (with the three darkest colours signalling dehydration, and the two middle colours underhydration; [3]). Values were recoded such that higher values indicated better hydration.

**Fig. 2.**
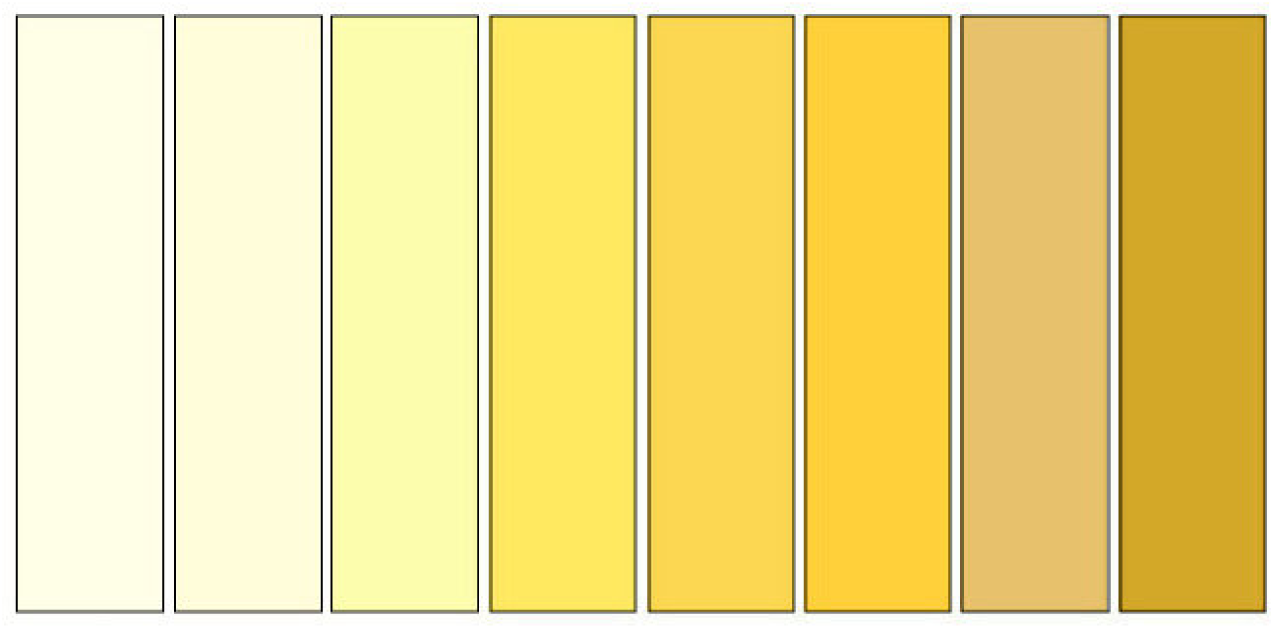
Eight-colour urine chart adapted from Armstrong et al. (1994)

We assessed void frequency as a secondary measure of hydration status by asking participants “How many times a day did you urinate yesterday?” A healthy void frequency for adults is at least seven times a day [3,7]. Participants also rated their confidence in their responses (0–100%). Urine colour and void frequency were correlated (*r* = 0.19, *p* < .001).

#### Socioeconomic status

Our main indicator of SES was educational attainment, as recorded by Prolific in six categories: no formal qualifications, secondary education (e.g., GED/GCSEs), high school diploma/A-levels, technical/community college, undergraduate degree (e.g., BA/BSc/other), graduate and/or doctorate degree (e.g., MA/MSc, Mphil). These categories were collapsed into two education levels: low (no formal qualifications, secondary education, high school diploma/A-levels) and high (technical/community college, undergraduate degree, graduate degree/doctorate).

Participants also reported their pretax household income by selecting one of five categories: less than £20,000, between £20,000 and £39,999, between £40,000 and £59,999, between £60,000 and £99,999, and more than £100,000. Education and household income were correlated (*r* = 0.26, *p* < .001).

#### Drink liking

Participants rated how much they liked various water types (tap water, water from cooler/fountain, bottled still water, bottled carbonated water, flavoured still bottled water, flavoured carbonated bottled water) and SSB types (carbonated soft drinks, non-carbonated sweet drinks, iced tea, other [ready-to-drink] sweetened beverages) on a scale from 0 = *not at all*, to 100 = *very much*. Overall water and SSB liking scores were computed by averaging the ratings across the specific drinks.

#### Control variables

##### Variables that affect hydration status

We assessed variables that may affect urine colour or void frequency: chronic health conditions (diabetes insipidus, renal disease, liver disease, gastrointestinal diseases or problems, cardiac or pulmonary disease), medication use (medications containing diuretics, phenytoin, lithium, demeclocycline, or amphotericin), dietary factors (severe salt restriction due to hypertension, high-protein diet, hypocaloric diet), and other factors (urinary tract infection, menstruation, pregnancy and/or lactation). Participants also reported whether they were colour blind given that this condition can affect perceptions of the urine colour scale.

##### Lifestyle variables

Participants additionally reported the number of portions of vegetables and fruit consumed on the previous day (based on NHS portion sizes; [20]). A modified version of the International Physical Activity Questionnaire (IPAC; [21]) was used to assess the number of minutes the previous day spent walking, in moderate activity, and in vigorous activity. These were converted into metabolic equivalent of task (MET) scores. Body Mass Index (BMI; kg/m^2^) was calculated from participants’ self-reported height and weight.

#### Knowledge and beliefs about urine colour

We asked open questions about participants’ beliefs regarding urine colour as a hydration marker: “What do you think the colour of someone’s urine reflects about how much they drink?”, “How do you think the colour of someone’s urine is related to their health?”, and “How does observing your urine colour affect your drinking behaviour, if it does at all?”. We coded responses to these questions to assess participants’ knowledge and reliance on their urine colour to gauge hydration needs and status (detailed coding information can be found in our online repository). Participants also indicated whether they had previously seen the urine colour chart (yes/no) and which colour(s) they think reflects healthy hydration.

Finally, participants provided demographic information such as age, gender, and current country of residence within the UK, and were then debriefed.

### Analysis plan

We tested our preregistered hypotheses using stepwise binomial logistic regression models predicting hydration status. We first regressed hydration status on water intake (H1) and SSB intake (H2) in Step 1, adding control variables that may affect hydration status in Step 2, and demographics and lifestyle variables in Step 3. To assess the association with SES (H3), we regressed hydration status on education level using the same stepwise approach.

We tested for robustness by (i) adjusting for intake of other beverage types, water from foods (portions of fruits and vegetables consumed), sugar added to drinks, and participant’s confidence in their urine-colour ratings); (ii) using void frequency (≥ 7 vs. < 7) as an alternative measure of hydration status; (iii) restricting analyses to the first assessment day to address day/weather effects; (iv) restricting analyses to participants reporting ≥80% confidence in their urine-colour rating, and (v) using multiple linear regression models with continuous urine colour as the outcome (see Table S1 in the SOM). For H3, we additionally examined (vi) education as categorical predictor, comparing each category to the lowest one (no formal qualifications); (vii) the same model as in “(vi)” but excluding the lowest category due to its small sample size; and (viii) models using household income (low; < £40,000 vs. high; ≥ £40,000) as an alternative proxy of SES (see Table S2 in the SOM). All analyses were conducted in *R* [22].

We used binomial logistic regression instead of the preregistered linear regression because diagnostic checks indicated violations of linearity assumptions (see SOM for details). However, linear models produced substantively similar results for all hypotheses (see Table S1 in the SOM).

## Results

### Descriptives

#### Prevalence of underhydration

Hydration status, as assessed with urine colour, indicated that 60% of participants were adequately hydrated, with urine colours in the three lightest categories. This pattern was consistent across females and males^2^ (see Fig. 3A). Of the 40% of participants who were inadequately hydrated, 35% were underhydrated and 5% were dehydrated (Perrier et al., 2021). Mean void frequency was 6.76 times per day (*SD* = 1.93), with those classified as adequately hydrated according to urine colour reporting a mean void frequency of 7.00 times (*SD* = 1.86), relative to 6.41 times (*SD* = 1.97) for those classified as inadequately hydrated.

**Fig. 3.**
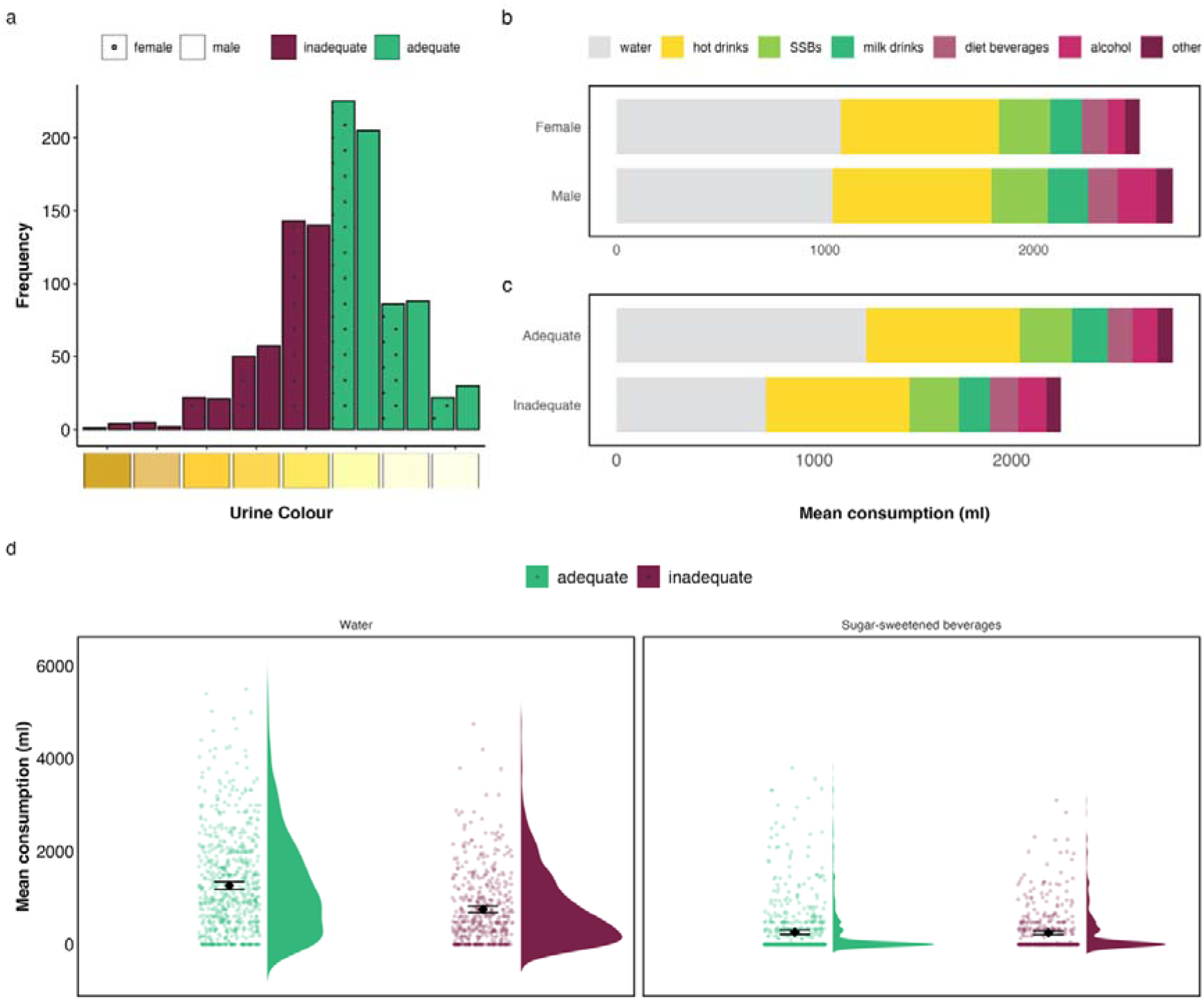
(a) Frequency of reported urine colour by gender and hydration status (inadequate vs. adequate) (b) Mean consumption of drink types by gender (c) Mean consumption of drink types by hydration status (d) Mean consumption of water and sugar-sweetened beverages (SSBs) by hydration status. Violin plots show the distributions; points and error bars represent means and 95% confidence intervals.

#### General fluid intake and gender differences

Mean total fluid intake (from drinks, milk in cereal, and soup, but excluding water from other foods) was 2.64 litres per day (*SD* = 1.09). Sixty-seven percent of females met the recommended minimum intake of 2 litres per day (*M* = 2.56, *SD* = 1.06), and 53% of males met the recommendation of 2.5 litres per day (*M* = 2.72, *SD* = 1.10; see Fig. 3B).

Water comprised the largest share of fluid intake: On average, participants consumed 1.06 litres of water per day (*SD* = 0.99), representing 37% of total intake with 76% of consumed water being tap water. Notably, 16% of participants reported drinking no water at all. SSBs accounted for 10% of total fluid intake, with a mean of 0.26 litres per day (*SD* = 0.53). Of the SSBs consumed, 40% were carbonated soft drinks (e.g., Coca-Cola and Fanta), 39% other [ready-to drink] sweet drinks (e.g., squash), and 19% were non-carbonated sweet drinks, and 2% were iced tea. Sixty-six percent of participants consumed no SSBs at all, 18% less than 500 ml, and fewer than 1% more than 1 litre per day.

#### Hydration status and fluid intake

There were observable differences in fluid intake between adequately and inadequately hydrated participants: On average, those classified as inadequately hydrated consumed 2.31 litres per day (*SD* = 0.98), including 0.76 litres of water (*SD* = 0.77), whereas adequately hydrated participants consumed 2.87 litres per day (*SD* = 1.09), including 1.26 litres of water (*SD* = 1.07). By contrast, SSB consumption varied little by hydration status, with inadequately hydrated participants reporting 0.25 litres per day (*SD* = 0.47) and adequately hydrated participants 0.26 litres per day (*SD* = 0.58) (see Fig. 3C and Fig. 3D).

#### SES and fluid intake

Participants with higher levels of education drank, on average, around 100 ml more fluid in total (*M* = 2.70, *SD* = 1.08) than those with lower levels of education (*M* = 2.56, *SD* = 1.09). The amount of water and SSBs consumed were very similar across education level: Participants with lower levels of education consumed, on average, 1 litre (*SD* = 0.98) of water and 0.27 litres (*SD* = 0.53) of SSBs per day, compared to 1.10 litre of water (*SD* = 1.00) and 0.25 litres (*SD* = 0.54) of SSBs per day among those with higher levels of education (see Fig. 4B).

**Fig. 4.**
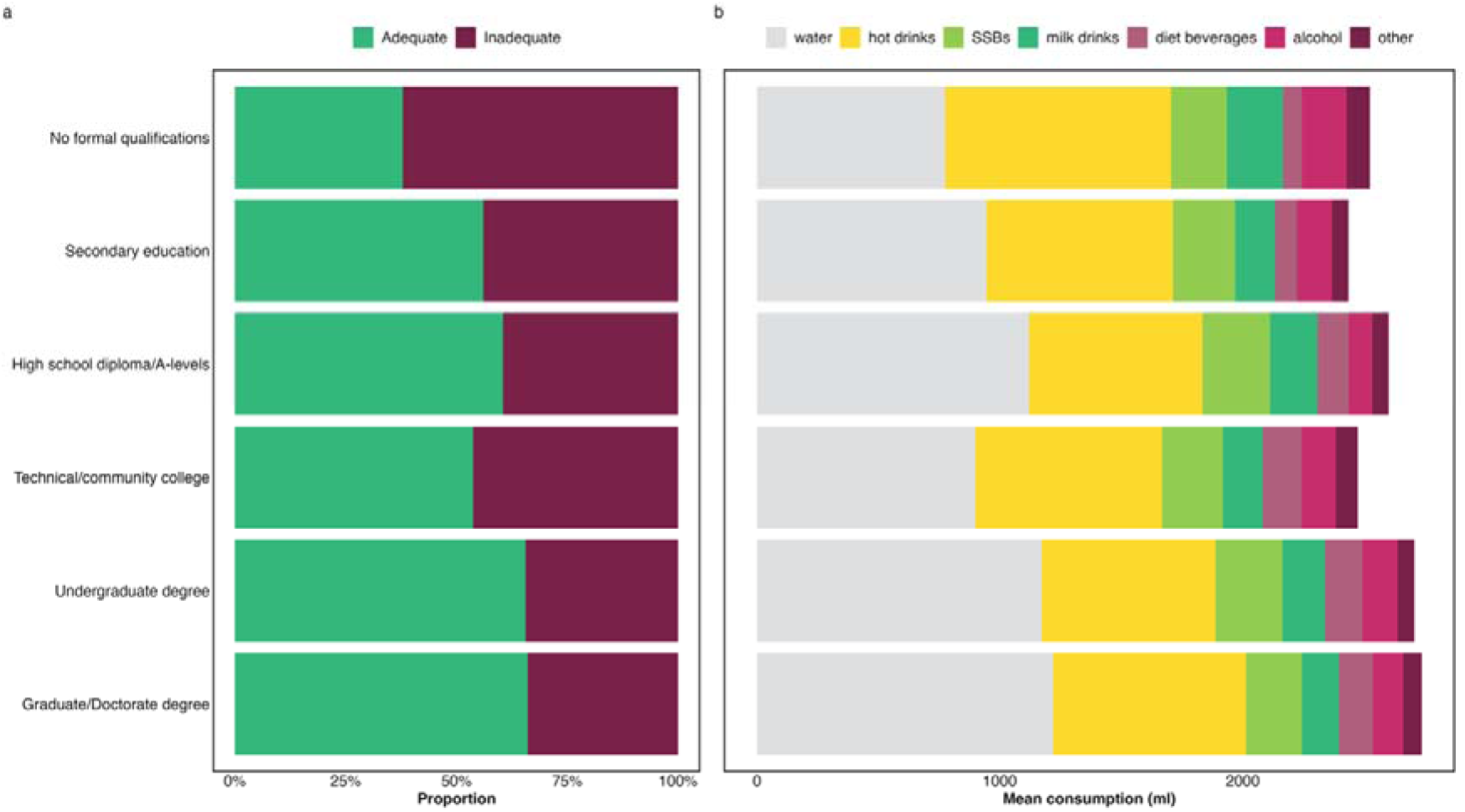
(a) Hydration status by education level (b) Mean beverage consumption by education level. *Note*. The “No formal qualifications” category consisted of only a small number of participants (*n* = 34); findings for this group should therefore be interpreted with caution.

### Confirmatory analyses

#### Association between water intake and hydration status (H1)

In line with our hypothesis, higher water intake was associated with a greater likelihood of being adequately hydrated, as assessed with urine colour (OR = 1.83, *p* < .001). This association held after adjustment for control variables (Step 2) and for demographic and lifestyle variables (Step 3; see Table 2), as well as in robustness checks with additional control variables (i.e., intake of other drinks, portions of fruits and vegetables consumed, sugar added to drinks, and participant’s confidence in their urine-colour ratings; see Table S1 in the SOM; see Fig. 3D). The findings suggest that drinking one extra glass of water (250 ml) per day increased the odds of being adequately hydrated by about 16%; drinking two extra glasses (500 ml) increased the odds by about 36%.

**Table 2.** Association between water intake and hydration status (adequate vs. inadequate)

| Variable | Step 1 |  |  | Step 2 |  |  | Step 3 |  |  |
| --- | --- | --- | --- | --- | --- | --- | --- | --- | --- |
|  | OR | 95% CI | p-value | OR | 95% CI | p-value | OR | 95% CI | p-value |
| Total water intake | 1.83 | 1.58, 2.13 | < .001 | 1.85 | 1.60, 2.16 | < .001 | 1.88 | 1.62, 2.20 | < .001 |
| Controls |  | No |  |  | Yes |  |  | Yes |  |
| Demographic and lifestyle variables |  | No |  |  | No |  |  | Yes |  |
| Observations |  | 1112 |  |  | 1112 |  |  | 1099 |  |
| Nagelkerke $R^2$ | | 0.09 | | | 0.10 | | | 0.12 | |

*Note.* OR = Odds Ratio, CI = Confidence Interval. Control variables included chronic health conditions, medication use, dietary factors, menstruation, pregnancy/lactation, colour blindness, and survey day and location. Demographic variables included age and gender; lifestyle variables included BMI and physical activity.

### Association between SSB intake and hydration status (H2)

Contrary to our hypothesis, initial analyses showed no association between SSB intake and the likelihood of being adequately hydrated (OR = 1.03, *p* = .686). This null effect remained after adjustment for control variables (Step 2), and for demographic and lifestyle variables (Step 3; see Table 3 and Fig. 3D). However, in robustness checks adjusting for additional control variables (i.e., intake of other drinks, portions of fruits and vegetables consumed, sugar added to drinks, and participant’s confidence in their urine-colour ratings) the association between SSB and hydration status became significant (OR = 1.46, *p* < .001; see Table S1 in the SOM). This pattern suggests a suppression effect: SSB intake was negatively correlated with both water intake (*r* = −0.22, *p* < .001) and total fluid intake (*r* = −0.53, *p* < .001), indicating that people who drink more SSBs consume less water and less fluid overall. Thus, SSBs appear to contribute to hydration only in the context of sufficient overall fluid intake^3^.

**Table 3.** Association between SSB intake and hydration status (adequate vs. inadequate)

| Variable | Step 1 |  |  | Step 2 |  |  | Step 3 |  |  |
| --- | --- | --- | --- | --- | --- | --- | --- | --- | --- |
|  | OR | 95% CI | p-value | OR | 95% CI | p-value | OR | 95% CI | p-value |
| Total SSB intake | 1.03 | 0.91, 1.16 | .686 | 1.03 | 0.91, 1.16 | .664 | 1.04 | 0.92, 1.19 | .503 |
| Controls |  | No |  |  | Yes |  |  | Yes |  |
| Demographic and lifestyle variables |  | No |  |  | No |  |  | Yes |  |
| Observations |  | 1112 |  |  | 1112 |  |  | 1099 |  |
| Nagelkerke $R^2$ | | < 0.01 | | | 0.01 | | | 0.03 | |

*Note.* OR = Odds Ratio, CI = Confidence Interval. Control variables included chronic health conditions, medication use, dietary factors, menstruation, pregnancy/lactation, colour blindness, survey day and location. Demographic variables included age and gender; lifestyle
variables included BMI and physical activity.

### Association between SES and hydration status (H3)

Contrary to our hypothesis, the likelihood of being adequately hydrated did not differ significantly between participants with higher (62% hydrated) and lower levels of education (57% hydrated; OR = 1.24, *p* = .084; see Table 4). This null effect remained after adjusting for control variables (Step 2), and for demographic and lifestyle variables (Step 3), as well as in robustness checks with additional control variables (i.e., intake of other drinks, portions of fruits and vegetables consumed, sugar added to drinks, and participant’s confidence in their urine-colour ratings; see Table S1 in the SOM; see Fig. 4A).

**Table 4.** Association between education and hydration status (adequate vs. inadequate)

| Variable | Step 1 |  |  | Step 2 |  |  | Step 3 |  |  |
| --- | --- | --- | --- | --- | --- | --- | --- | --- | --- |
|  | OR | 95% CI | p-value | OR | 95% CI | p-value | OR | 95% CI | p-value |
| Education |  |  |  |  |  |  |  |  |  |
| Low | — | — |  | — | — |  | — | — |  |
| High | 1.24 | 0.97, 1.58 | .084 | 1.26 | 0.99, 1.62 | .065 | 1.22 | 0.95, 1.57 | .122 |
| Controls |  | No |  |  | Yes |  |  | Yes |  |
| Demographic and lifestyle variables |  | No |  |  | No |  |  | Yes |  |
| Observations |  | 1112 |  |  | 1112 |  |  | 1099 |  |
| Nagelkerke $R^2$ | | < 0.01 | | | 0.02 | | | 0.03 | |

*Note.* OR = Odds Ratio, CI = Confidence Interval. Control variables included chronic health conditions, medication use, dietary factors, menstruation, pregnancy/lactation, colour blindness, and survey day and location. Demographic variables included age and gender; lifestyle variables included BMI and physical activity.

However, in a further set of robustness checks, SES was associated with the likelihood of being adequately hydrated when alternative indicators were used, such as household income or more fine-grained comparisons across education levels. Specifically, comparing across all education categories showed that participants with a higher education level—particularly those with undergraduate or postgraduate degrees—were more likely to be adequately hydrated than those with the lowest education levels (see Table S2 in SOM).

All findings for all hypotheses were robust across our robustness checks; see Table S1 in the SOM.

### Exploratory analyses

#### Correlations between drink liking, intake, and hydration status

Water liking was positively associated with water intake (*r* = 0.19, *p* < .001) but not hydration status. SSB liking was positively associated with SSB intake (*r* = 0.29, *p* < .001) but negatively with water intake (*r* = –0.06, *p* = .047) and hydration status (*r* = –0.08, *p* = .005). In addition, water and SSB liking were positively associated (*r* = 0.37, *p* < .001).

#### Knowledge and beliefs about urine colour

Participants’ open-ended responses indicated that while the majority (89%) associated urine colour with fluid consumption and/or hydration status, fewer reported acting on that information: Around 50% said they drank more after observing darker urine, but about 25% indicated that urine colour does not influence their drinking behaviour. Among those who associated urine colour with hydration, adequately hydrated participants were more likely to report drinking more after observing darker urine (53%) than inadequately hydrated participants (43%). Nonetheless, there was a notable knowledge–behaviour gap in both hydration groups. Finally, 42% of participants reported having seen the urine colour chart before, but only 26% correctly identified the colours indicating a healthy hydration status.

## Discussion

The results of this cross-sectional study in a large UK sample provides insights into fluid intake patterns, hydration status, and their relation to education level. Our preregistered analyses showed that higher water intake was associated with greater likelihood of adequate hydration. In contrast, SSB intake showed no overall association with hydration status unless intake from other drink sources was included in the model, suggesting that individuals who consume SSBs consume fewer total fluids and are hence less hydrated. Indeed, individuals who were inadequately hydrated did not consume less SSBs than those who were adequately hydrated—they consumed less water. This suggests that water intake, rather than SSB intake, distinguishes adequately from inadequately hydrated participants.

Regarding SES, education level (dichotomised) was not significantly associated with hydration status. Finer-grained comparisons across all education levels revealed that participants with higher education levels were better hydrated, but the pattern was inconsistent (i.e., there was no significant effect for participants in the category “Technical/community college”; see Fig. 4A). In contrast, household income was significantly associated with hydration status. This divergence may reflect that income more directly captures material resources and daily circumstances relevant to hydration behaviour—such as the affordability and availability of beverages or workplace conditions that limit drinking—whereas education alone may not consistently translate into such practical differences [23].

Beyond these confirmatory findings, several descriptive patterns provide context for hydration behaviours in this UK sample. Although mean total fluid intake was broadly in line with recommendations for men and women, a substantial proportion of participants did not meet recommended water-intake guidelines. Indeed, self-reported urine colour suggested that 40% of participants were inadequately hydrated (of which 5% were dehydrated) on the previous day. This finding is consistent with previous research showing that around 30–40% of UK participants report urine colours indicative of underhydration [11,24], and that those who drink more water report significantly lighter urine [25].

While, on average, men consumed slightly more fluid overall than women, they were less likely to meet sex-specific recommendations. Although the average total fluid intake was adequate, water accounted for only 37% of total intake, and 16% of participants reported consuming no water at all. This indicates that many people rely on beverages other than plain water---most notably hot drinks---as their primary source of fluids. From a public health perspective, this is relevant because consumption of hot drinks was positively correlated with amount of added sugar in drinks (*r* = 0.27, *p* < .001), and consumption of sugar-sweetened beverages is associated with adverse health outcomes such as obesity and dental caries [26,27].

The relatively modest effect of education suggests that factors beyond educational background—such as individual preferences or environmental access to palatable water—may play a role in shaping hydration patterns. Consistent with this interpretation, our exploratory analyses showed that liking for water was positively associated with water intake, while liking for SSBs was positively associated with greater SSB intake and worse hydration. These preference–intake links suggest potential behaviour pathways that may be leveraged in interventions.

### Implications and future directions

The findings of this study carry important public health implications. Underhydration—known to have significant implications for physical and cognitive and functioning [1–3]—was common in our sample (40%). Preventing and addressing inadequate hydration should be considered a public health priority. Because water intake is a complex health behaviour shaped by numerous individual and situational determinants [28], effective approaches will likely require complex interventions that involve multiple individual and system-level stakeholders [29]. One approach is to increase the attractiveness of water and reduce the appeal of SSBs—for example, through advertising or labelling that highlights the immediate rewards of drinking water, such as its refreshing taste and ability to quench thirst [30,31]. Further research could test whether reducing the appeal of SSBs can indirectly promote better hydration by reducing their consumption and encouraging water intake [32,33].

Moreover, hydration could be supported by improving public knowledge and use of urine colour as a simple self-monitoring tool: Only half of our participants reported adjusting their drinking in response to urine colour, and only 26% could correctly identify the colours that indicate healthy hydration. To increase awareness and usability, urine colour charts could be integrated into school curricula al alongside dietary guidelines, displayed in general practitioners’ offices, or placed in highly visible locations such as toilet stalls in hospitals and other public facilities. Beyond raising awareness, such cues may also function as actionable prompts for behaviour change, particularly given our finding that even individuals who have awareness do not translate that into increased drinking when their urine colour is dark. Future research should examine how these approaches can effectively increase water intake and improve hydration status, and help bridge the gap between knowledge and action.

In addition, policy measures may also help to promote adequate hydration. These are likely to be most effective when they focus on increasing the accessibility in public settings and promoting water as the primary beverage choice in schools, hospitals, prisons, public buildings, public transport settings, and other public institutions. Because toilet availability has been identified as a barrier to drinking water while travelling, improving access to safe and clean toilet facilities in public transport and other settings could further help prevent underhydration [28]. Finally, price-based measures may also play a role, such as raising sugar taxes to discourage sugar-sweetened beverage consumption and removing value-added tax (sales tax) on bottled water [34].

### Strengths and limitations

To our knowledge, this is the first UK-based study to go beyond analyses of total fluid intake, and examine hydration status in relation to intake of specific beverage types and socioeconomic differences.

One limitation is the reliance on self-reported urine colour rather than physiological markers; however, findings were consistent when using void frequency as a secondary measure of hydration status in robustness analyses. In addition, urine colour has been validated as an effective indicator of hydration in laboratory settings [34,35] and relates to urine concentration, unlike self-reported thirst levels [36], which are easily overruled, ignored, or forgotten [24].

Another limitation is the 24-hour fluid intake assessment, which may underestimate total fluid intake relative to 7-day fluid records [37]. This approach was chosen to achieve balanced educational representation given lower availability and higher dropout among low-SES participants in longer assessments [38]. Nevertheless, our results aligned relatively well with other findings: The percentage of women and men adhering to EFSA fluid recommendations in our sample was comparable to that reported by Ferreira-Pêgo et al. [5]^4^ but lower than that reported by Gibson and Shirreffs [39]. Consistent with the latter study, we found that hot drinks were the most popular beverage, with the highest percentage of the sample reporting consumption, closely followed by water (see Fig. S2 in SOM). In contrast, a smaller percentage of participants reported consuming milk drinks and alcohol. This may reflect our assessment of intake on a single weekday, whereas Gibson and Shirreffs [39] assessed consumption over an entire week, including weekend days.

### Conclusion

Our results emphasise the importance of water intake for adequate hydration, and indicate modest SES-related differences that merit further investigation. Interventions that increase the appeal and availability of water—and that remove barriers to drinking water—may improve hydration and help reduce inequalities in hydration status. Complementary strategies to reduce SSB consumption remain important for broader public health reasons. Future work would benefit from employing longitudinal or experimental designs, multiple-day intake assessments, and objective biomarkers to clarify causal mechanisms and test targeted interventions.

## Declarations

### Author contributions

M.A.C. and E.K.P. conceptualised the study. M.A.C. collected and curated data, conducted the analyses, and wrote the original draft of the manuscript. M.A.C. and E.K.P. reviewed and edited the manuscript.

### Ethics statement

The research protocol was approved by the College of Science and Engineering Ethics Committee, University of Glasgow (application number: 200210197). All methods were carried out in accordance with relevant guidelines and regulations. Written informed consent was obtained from participants prior to participation.

## Acknowledgments

We thank Susannah Goss for scientific editing, Sevasti Karagianni for assistance with coding the open-ended responses, and Piero Ronzani for advice on the figures.

## Data availability

The data supporting this study’s findings are available on: https://osf.io/jrwyh/overview?view_only=b51a7b252adc46288a27f3ed46c11c18 and https://github.com/maclaassen/hydration.

## Footnotes

1 Hydration status correlated positively with the number of glasses of water consumed, *r* = 0.30, *p* < .001 (Survey 1) and *r* = 0.22, *p* = .006 (Survey 2), and negatively with the number of glasses of SSBs consumed: *r* = −0.27, *p* < .001 (Survey 1) and *r* = −0.21, *p* = .009 (Survey 2).

2 Because less than 1% of our sample identified as non-binary or other, we only compared female and male participants.

3 Given that hot drinks were the most commonly consumed beverage in our sample, we explored their association with hydration status. Hot drink intake was not associated with hydration status when modelled alone (OR = 1.08, p = .190), nor when included alongside SSB intake (OR = 1.09, p = .166). However, hot drink intake was positively associated with adequate hydration when modelled together with water intake (OR = 1.22, p = .003), suggesting a contribution to hydration once water intake is accounted for. Similar to SSB intake, hot drink intake was negatively correlated with water intake (r = −0.186, p < .001), indicating that individuals may substitute hot drinks for water. Also, hot drink intake was positively correlated with added sugar to drinks (r = 0.27, p < .001). Importantly, including hot drinks in the models does not alter the central conclusion that water intake—rather than SSB intake—is associated with adequate hydration.

4 They set slightly lower reference values because they subtracted 20% for food-sourced fluids.

## References

1. Dmitrieva, N. I., Boehm, M., Yancey, P. H., & Enhörning, S. (2024). Long-term health outcomes associated with hydration status. Nature Reviews Nephrology, 20(5), 275–294. 10.1038/s41581-024-00817-1

2. Liska, D., Mah, E., Brisbois, T., Barrios, P. L., Baker, L. B., & Spriet, L. L. (2019). Narrative review of hydration and selected health outcomes in the general population. Nutrients, 11(70). 10.3390/nu11010070

3. Perrier, E. T., Armstrong, L. E., Bottin, J. H., Clark, W. F., Dolci, A., Guelinckx, I., Iroz, A., Kavouras, S. A., Lang, F., Lieberman, H. R., Melander, O., Morin, C., Seksek, I., Stookey, J. D., Tack, I., Vanhaecke, T., Vecchio, M., & Péronnet, F. (2021). Hydration for health hypothesis: A narrative review of supporting evidence. European Journal of Nutrition, 60(3), 1167–1180. 10.1007/s00394-020-02296-z

4. EFSA. (2010). Scientific opinion on dietary reference values for water. EFSA Journal, 8(3), 1459. 10.2903/j.efsa.2010.1459

5. Ferreira-Pêgo, C., Guelinckx, I., Moreno, L. A., Kavouras, S. A., Gandy, J., Martinez, H., Bardosono, S., Abdollahi, M., Nasseri, E., Jarosz, A., Babio, N., & Salas-Salvadó, J. (2015). Total fluid intake and its determinants: Cross-sectional surveys among adults in 13 countries worldwide. European Journal of Nutrition, 54(S2), 35–43. 10.1007/s00394-015-0943-9

6. Malisova, O., Athanasatou, A., Pepa, A., Husemann, M., Domnik, K., Braun, H., Mora-Rodriguez, R., Ortega, J. F., Fernandez-Elias, V. E., & Kapsokefalou, M. (2016). Water intake and hydration indices in healthy European adults: The European Hydration Research Study (EHRS). Nutrients, 8(4). 10.3390/nu8040204

7. Adams, J. D., Arnaoutis, G., Johnson, E. C., Jansen, L. T., Bougatsas, D., Capitan-Jimenez, C., Mauromoustakos, A., Panagiotakos, D. B., Perrier, E. T., Guelinckx, I., & Kavouras, S. A. (2021). Combining urine color and void number to assess hydration in adults and children. European Journal of Clinical Nutrition, 75(8), 1262–1266. 10.1038/s41430-020-00834-w

8. Perrier, E. T., Johnson, E. C., McKenzie, A. L., Ellis, L. A., & Armstrong, L. E. (2016). Urine colour change as an indicator of change in daily water intake: A quantitative analysis. European Journal of Nutrition, 55(5), 1943–1949. 10.1007/s00394-015-1010-2

9. NHS. (2025, September). Hydration: Staying hydrated is easy when you know how. NHS.Uk. https://www.nhsinform.scot/campaigns/hydration/

10. Maughan, R. J., Watson, P., Cordery, P. A., Walsh, N. P., Oliver, S. J., Dolci, A., Rodriguez-Sanchez, N., & Galloway, S. D. (2016). A randomized trial to assess the potential of different beverages to affect hydration status: Development of a beverage hydration index. The American Journal of Clinical Nutrition, 103(3), 717–723. 10.3945/ajcn.115.114769

11. Claassen, M. A., Lomann, M., & Papies, E. K. (2023). Increased consumption despite fewer occasions: A longitudinal analysis of COVID-19 lockdown effects on soft drink consumption in England. Appetite, 187(106579). 10.1016/j.appet.2023.106579

12. Bougatsas, D., Arnaoutis, G., Panagiotakos, D. B., Seal, A. D., Johnson, E. C., Bottin, J. H., Tsipouridi, S., & Kavouras, S. A. (2018). Fluid consumption pattern and hydration among 8–14 years-old children. European Journal of Clinical Nutrition, 72, 420–427. 10.1038/s41430-017-0012-y

13. Illescas-Zarate, D., Espinosa-Montero, J., Flores, M., & Barquera, S. (2015). Plain water consumption is associated with lower intake of caloric beverage: Cross-sectional study in Mexican adults with low socioeconomic status. BMC Public Health, 15(405). 10.1186/s12889-015-1699-0

14. Basu, S., McKee, M., Galea, G., & Stuckler, D. (2013). Relationship of soft drink consumption to global overweight, obesity, and diabetes: A cross-national analysis of 75 countries. American Journal of Public Health, 103(11), 2071–2077. 10.2105/AJPH.2012.300974

15. Brooks, C. J., Gortmaker, S. L., Long, M. W., Cradock, A. L., & Kenney, E. L. (2017). Racial/ethnic and socioeconomic disparities in hydration status among US adults and the role of tap water and other beverage intake. American Journal of Public Health, 107(9), 1387–1394. 10.2105/AJPH.2017.303923

16. Bolt-Evensen, K., Vik, F. N., Stea, T. H., Klepp, K.-I., & Bere, E. (2018). Consumption of sugar-sweetened beverages and artificially sweetened beverages from childhood to adulthood in relation to socioeconomic status—15 years follow-up in Norway. International Journal of Behavioral Nutrition and Physical Activity, 15(8). 10.1186/s12966-018-0646-8

17. Prolific. (2014). Prolific.Co. https://www.prolific.co

18. Johnson, E. C., Péronnet, F., Jansen, L. T., Capitan-Jiménez, C., Adams, J., Guelinckx, I., Jiménez, L., Mauromoustakos, A., & Kavouras, S. A. (2017). Validation testing demonstrates efficacy of a 7-day fluid record to estimate daily water intake in adult men and women when compared with total body water turnover measurement. The Journal of Nutrition, 147(10), 2001–2007. 10.3945/jn.117.253377

19. Armstrong, L. E. (1994). Urinary indices of hydration status. International Journal of Sport Nutrition, 4, 265–279. 10.1123/ijsn.4.3.265

20. NHS. (2022, July). 5 A Day portion sizes. NHS.Uk. https://www.nhs.uk/live-well/eat-well/5-a-day/portion-sizes/

21. Craig, C. L., Marshall, A. L., Sjöström, M., Bauman, A. E., Booth, M. L., Ainsworth, B. E., Pratt, M., Ekelund, U., Yngve, A., Sallis, J. F., & Oja, P. (2003). International physical activity questionnaire: 12-country reliability and validity. Medicine and Science in Sports and Exercise, 35(8), 1381–1395. 10.1249/01.mss.0000078924.61453.fb

22. R Core Team. (2020). R: A language and environment for statistical computing [Computer software]. https://www.R-project.org/

23. Claassen, M. A., Corneille, O., & Klein, O. (2019). Psychological consequences of inequality for food intake. In J. Jetten & K. Peters (Eds), The Social Psychology of Inequality (pp. 155–172). Springer. 10.1007/978-3-030-28856-3_10

24. Rodger, A., Wehbe, L. H., & Papies, E. K. (2021). “I know it’s just pouring it from the tap, but it’s not easy”: Motivational processes that underlie water drinking. Appetite, 164(105249). 10.1016/j.appet.2021.105249

25. Rodger, A., & Papies, E. K. (2022). “I don’t just drink water for the sake of it”: Understanding the influence of value, reward, self-identity and early life on water drinking behaviour. Food Quality and Preference, 99(104576). 10.1016/j.foodqual.2022.104576

26. Hasselkvist, A., Johansson, A., & Johansson, A.-K. (2014). Association between soft drink consumption, oral health and some lifestyle factors in Swedish adolescents. Acta Odontologica Scandinavica, 72(8), 1039–1046. 10.3109/00016357.2014.946964

27. Schulze, M. B., Manson, J. E., Ludwig, D. S., Colditz, G. A., Stampfer, M. J., Willett, W. C., & Hu, F. B. (2004). Sugar-sweetened beverages, weight gain, and incidence of type 2 diabetes in young and middle-aged women. JAMA, 292(8), 927–934. 10.1001/jama.292.8.927

28. Rodger, A., Barsalou, L. W., & Papies, E. K. (2024). Habitualness, reward and external constraints: Exploring the underlying influences of daily water intake using the Situated Assessment Method^2^. Applied Psychology: Health and Well-Being, 16(4), 2458–2483. 10.1111/aphw.12598

29. Skivington, K., Matthews, L., Simpson, S. A., Craig, P., Baird, J., Blazeby, J. M., Boyd, K. A., Craig, N., French, D. P., McIntosh, E., Petticrew, M., Rycroft-Malone, J., White, M., & Moore, L. (2021). A new framework for developing and evaluating complex interventions: Update of Medical Research Council guidance. BMJ, 374(n2061). 10.1136/bmj.n2061

30. Claassen, M. A., Rusz, D., & Papies, E. K. (2022). No evidence that consumption and reward words on labels increase the appeal of bottled water. Food Quality and Preference, 96(104403). 10.1016/j.foodqual.2021.104403

31. Claassen, M. A., & Papies, E. K. (2023). Representational shifts: Increasing motivation for bottled water through simulation-enhancing advertisements. BMC Public Health, 23(2209). 10.1186/s12889-023-17109-1

32. Papies, E. K., Claassen, M. A., Rusz, D., & Best, M. (2022). Flavors of desire: Cognitive representations of appetitive stimuli and their motivational implications. Journal of Experimental Psychology: General, 151(8), 1919–1941. 10.1037/xge0001157

33. Von Philipsborn, P., Stratil, J. M., Burns, J., Busert, L. K., Pfadenhauer, L. M., Polus, S., Holzapfel, C., Hauner, H., & Rehfuess, E. (2019). Environmental interventions to reduce the consumption of sugar-sweetened beverages and their effects on health. Cochrane Database of Systematic Reviews, 2019(6). 10.1002/14651858.CD012292.pub2

34. Wardenaar, F. C., Thompsett, D., Vento, K. A., & Bacalzo, D. (2021). A lavatory urine color (LUC) chart method can identify hypohydration in a physically active population. European Journal of Nutrition, 60(5), 2795–2805. 10.1007/s00394-020-02460-5

35. Wardenaar, F. C., Thompsett, D., Vento, K. A., Pesek, K., & Bacalzo, D. (2021). Athletes’ self-assessment of urine color using two color charts to determine urine concentration. International Journal of Environmental Research and Public Health, 18(4126). 10.3390/ijerph18084126

36. Wardenaar, F. C., Whitenack, L., Vento, K. A., Seltzer, R. G. N., Siegler, J., & Kavouras, S. A. (2024). Validity of combined hydration self-assessment measurements to estimate a low vs. high urine concentration in a small sample of (tactical) athletes. European Journal of Nutrition, 63(1), 185–193. 10.1007/s00394-023-03254-1

37. Bardosono, S., Monrozier, R., Permadhi, I., Manikam, N. R. M., Pohan, R., & Guelinckx, I. (2015). Total fluid intake assessed with a 7-day fluid record versus a 24-h dietary recall: A crossover study in Indonesian adolescents and adults. European Journal of Nutrition, 54(S2), 17–25. 10.1007/s00394-015-0954-6

38. Schmidt, S. C. E., & Woll, A. (2017). Longitudinal drop-out and weighting against its bias. BMC Medical Research Methodology, 17(164). 10.1186/s12874-017-0446-x

39. Gibson, S., & Shirreffs, S. M. (2013). Beverage consumption habits “24/7” among British adults: Association with total water intake and energy intake. Nutrition Journal, 12(9). 10.1186/1475-2891-12-9

